# Facial basal cell carcinoma: a clinicopathological analysis of incomplete excision

**DOI:** 10.1101/2021.12.02.21267192

**Authors:** Thomas Layton

## Abstract

Basal cell carcinoma (BCC) is the most common malignancy in humans. Incomplete excision following conventional surgical excision requires careful consideration given the potential for disease recurrence. We performed a retrospective study analyzing facial BCC treated by conventional surgical excision to investigate factors influencing the likelihood of tumour clearance. In total, 456 cases of facial BCC were reviewed to collect a dataset of 50 tumours with involved margins and these compared to 50 completely excised tumours from the same cohort. Statistical comparison between incomplete and complete excision cases was performed using several metrics including tumour location, histology, grade, surgeon experience and method of wound reconstruction. Interestingly, our results demonstrated that only tumour location and histological type had a statistically significant impact on the completeness of resection. Infiltrative, morphoeic and mixed tumours had a higher chance of incomplete excision, as did tumours located on the inner canthus and ala nasi. In addition, the overall incomplete excision rate was 10.96% in line with previous studies. Our results help inform surgical practice and support consideration of extending the recommended macroscopic surgical margin for higher risk tumours. More research is needed to further categorize facial BCC to optimize surgical management.

## Introduction

Basal cell carcinoma (BCC) is the most common malignant neoplasm in humans^1,2^. BCC is an indolent and locally invasive cancer most prevalent in Caucasians with over 200,000 BCCs excised in the UK during 2010 and from 1999 to 2010 the numbers of surgically treated BCCs increased by 81%^3,4^. This data indicates that in the UK BCC is nearly as common as all other cancers combined^3^.

Disease recurrence is a feared clinical entity in any malignancy and in up to 2% of BCCs treated by conventional methods, either surgical excision or curettage, the tumour will recur at a later date^3^. However, following a confirmed complete excision the risk of recurrence is significantly reduced^5^. Reported rates of incomplete excision in BCC at all anatomical sites range from 4% to 16.6%^3,5^ and a comprehensive study analyzing peripheral and deep margins of 1539 BBCs excised by conventional surgery demonstrated an overall incomplete excision rate of ∼8%^3^.

Despite the importance of these studies, further classification of facial BCC is warranted as it represents a distinct clinicopathological entity. Indeed, surgical management of facial BCC mandates careful consideration due to the potential for severe disfigurement and the challenge in optimizing functional and cosmetic outcomes. In this study, we provide a valuable new cohort of facial BCC treated at a large tertiary centre and define distinct factors influencing successful excision. Our results may help to refine surgical practice and inform best clinical guidelines for this common malignancy.

## Materials and Methods

We performed a retrospective analysis of BCC treated between October 2013 and May 2014 at a large tertiary referral centre. Pathology reports were reviewed to categorize excision margins as complete or incomplete. Incomplete excision was taken as surgical margins less than 1 mm at either deep or local margin as defined by the British Association of Dermatologists (BAD) guidelines^6^. Patients were treated with conventional surgical excision using predefined margins according to BAD guidelines and all tumours were primary lesions without previous treatment.

Surgeons were either dermatologists, oral and maxillofacial surgeons or plastic surgeons. Specimens were oriented using a marker stitch and all diagnosis were confirmed by a dermatological histopathologist. Patients with BCC located outside of the facial region were excluded. We could then define the rate of incomplete excision of facial BCC.

A total of 456 patients were reviewed to collect a sample of 50 tumours with incomplete surgical margins. All lesions with incomplete margins were categorized with regards to tumour site, grade of tumour, tumour histology, grade of surgeon, reconstruction method and follow-up. Follow up was stratified into three categories: re-excision, radiotherapy or surveillance. Methods of reconstruction were primary closure, skin graft, or local or distant flap. A random sample of completely resected facial BCC was collected from the same time period using a random number generator.

In order to undertake a systematic investigation of tumour location the face was divided into 10 subunits: temple, forehead, scalp, periorbital region, inner canthus, ala nasi, nose (elsewhere), auricular, cheek and perioral region. Lesions in each subunit were evaluated according to the previously defined variables. Using 10 areas of the face may influence the significance of statistical comparisons, but one aim of our study was to investigate localised areas of the face at higher risk of incomplete excision. The cheek included any tumours lateral to the nasolabial fold, inferior to the infraorbital region and medial to the preauricular area

All data analysis was performed in RStudio (R Version 3.5) and *p*-values were calculated using Fisher’s Exact test. *p*-values < 0.05 were taken as significant. No methods were used to predefine sample sizes.

## Results

A retrospective analysis of 456 facial BCCs treated between October 2013 and May 2014 was undertaken to collect a sample of 50 incompletely excised tumours, with an overall incomplete excision rate of 10.96%. Tumours with involved margins included 16 (32%) female and 34 (68%) male patients. The mean age was 75.6 years (35-94 years). Of the complete excision tumours, 24 (48%) were female and 26 (52%) were male and the mean age was 72.45 years (41-92 years).

For incomplete tumours, 32% (16/50) were infiltrative, 32% (16/50) mixed, 28% (14/50) morphoeic, 4% (2/50) ulcerated, 2% (1/50) cystic and 2% superficial (1/50) variants. For complete excision tumours, 56% (27/50) were nodular, 16% (7/50) infiltrative, 14% (7/50) ulcerated, 2% (1/50) mixed, 10% (5/50) superficial, 2% (1/50) morphoeic and 4% (2/50) cystic variants (Table A). There was a statistically significant difference between nodular (*p*=0.0001), infiltrative (*p*=0.0279), morphoeic (*p*=0.0039) and mixed (*p*=0.0001) variants.

**Table A - B.** Location and histological variant of facial basal cell c arcinoma in incomplete and complete excision cohorts (*n* = 50).

| LOCATION | INCOMPLETE | COMPLETE | % INCOMPLETE | P-VALUE |
| --- | --- | --- | --- | --- |
| TEMPLE | 11 | 9 | 55 | 0.8031 |
| FOREHEAD | 1 | 5 | 29 | 0.2180 |
| SCALP | 1 | 1 | 50 | 1.0000 |
| PERIORBITAL | 1 | 0 | 100 | NA |
| INNER CANTHUS | 6 | 1 | 86 | 0.0500* |
| ALA NASI | 10 | 1 | 91 | 0.0039* |
| NOSE (ELSEWHERE) | 2 | 8 | 20 | 0.0458* |
| AURICULA | 10 | 8 | 55 | 0.3976 |
| CHEEK | 5 | 15 | 25 | 0.0001* |
| PERIORAL | 3 | 2 | 60 | 0.5000 |

| HISTOLOGY | INCOMPLETE | COMPLETE | % INCOMPLETE | P-VALUE |
| --- | --- | --- | --- | --- |
| CYSTIC | 1 | 2 | 33 | 0.5000 |
| NODULAR | 4 | 27 | 13 | 0.0001* |
| INFILTRATIVE | 16 | 7 | 69 | 0.0279* |
| MORPHOEIC | 10 | 1 | 91 | 0.0039* |
| SUPERFICIAL | 1 | 5 | 20 | 0.1022 |
| ULCERATED | 2 | 7 | 22 | 0.0798 |
| MIXED | 16 | 1 | 94 | 0.0001* |

In the incomplete excision tumours 22% (11/50) were located at the temple, 20% (10/50) the ala nasi, 20% (10/50) the auricular region, 12% (6/50) the inner canthus, 6% (3/50) the perioral region, 2% (1/50) the scalp, 4% (2/50) the nose (elsewhere), 2% (1/50) the periorbital region, 2% (1/50) the forehead and 10% (5/50) the cheek.

In the complete excision tumours 18% (9/50) were located at the temple, 2% (1/50) the ala nasi, 16% (8/50) the auricular region, 2% (1/50) the inner canthus, 4% (2/50) the perioral region, 2% (1/50) the scalp, 16% (8/50) the nose (elsewhere), 0% (0/50) the periorbital region, 10% (5/50) the forehead and 30% (15/50) the cheek. Tumours at the ala nasi (*p*=0.0039) and inner canthus (*p*=0.05) were significantly greater in incompletely excised tumours. In contrast, tumours at the cheek (*p*=0.0001) and nose outside of the alar region (*p*=0.0458) were significantly reduced in the incompletely excised tumours (Figure 1 & Table B).

**Figure 1:**
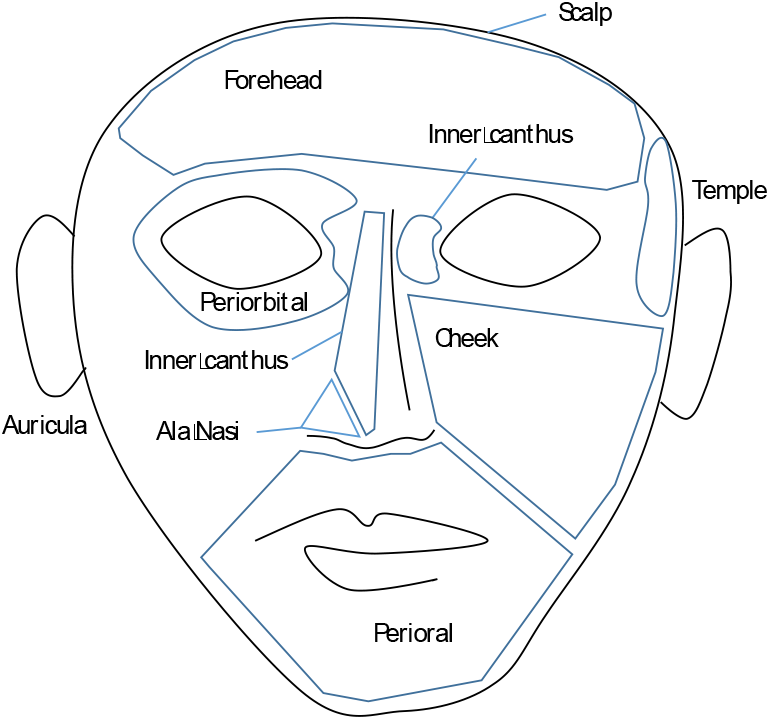
Schematic showing division of the facial into 10 sub regions.

All complete excision tumours were AJCC grade pT1 and none (0/50) exhibited perivascular or perineural infiltration. Of the incomplete excision tumours 90% (45/50) were pT1, 2% (1/50) were pT2 and 8% (4/50) were pT3. Of the incomplete excision tumours, 2% (1/50) exhibited perivascular infiltration, 2% (1/50) perineural infiltration and 2% (1/50) subcutaneous infiltration.

We next examined the involved margins for incomplete excision tumours. 40% (20/50) demonstrated an incomplete deep margin, 28% (14/50) an incomplete peripheral margin and 32% (16/50) had both incomplete deep and peripheral margins (Table C).

**Table C - F.**
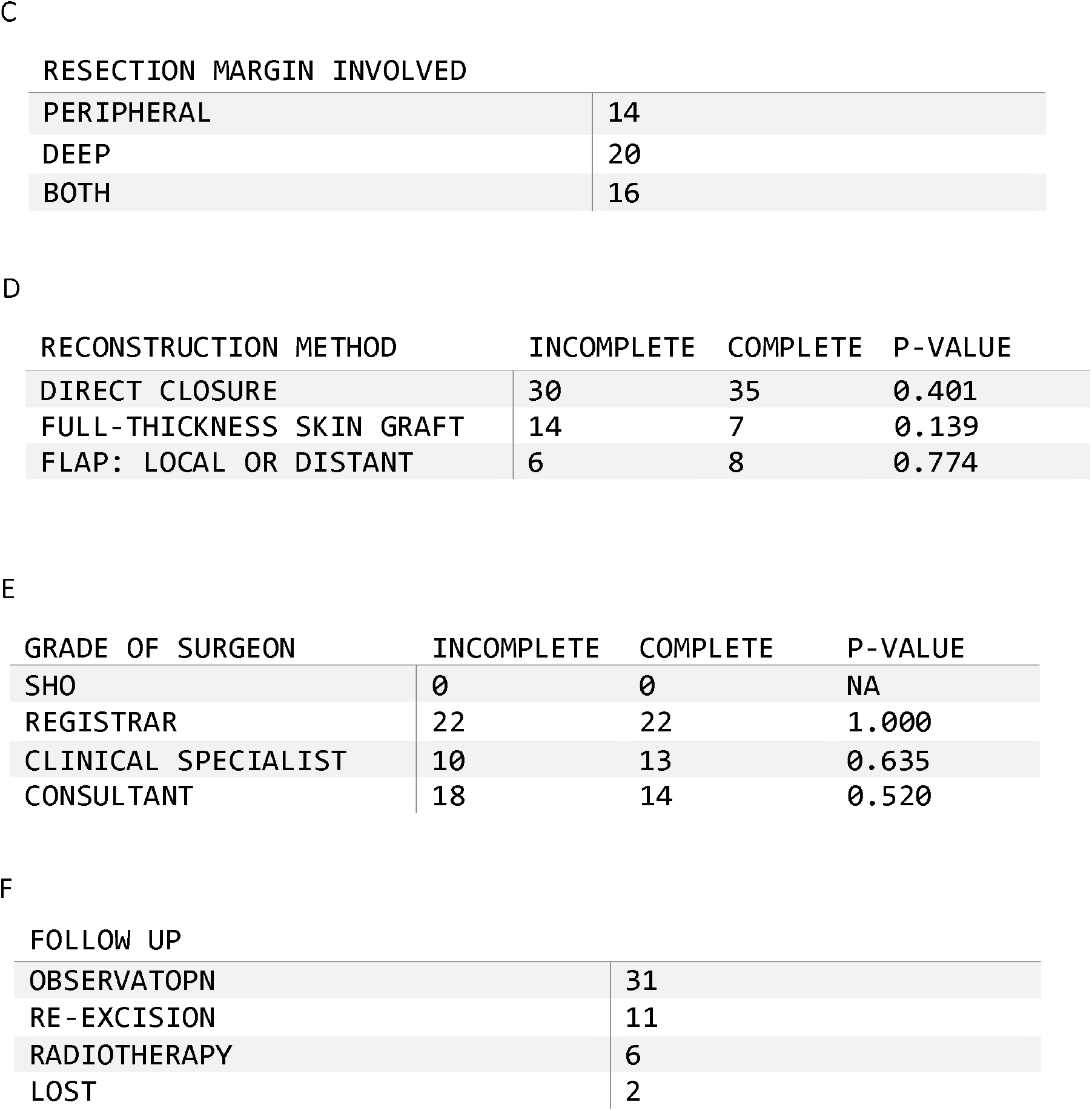
Reconstruction methods, grade of surgeon, resection margin involved and follow up of facial basal cell carcinoma in incomplete and complete excision cohorts (*n* = 50).

| RESECTION MARGIN INVOLVED |  |
| --- | --- |
| PERIPHERAL | 14 |
| DEEP | 20 |
| BOTH | 16 |

D
| RECONSTRUCTION METHOD | INCOMPLETE | COMPLETE | P-VALUE |
| --- | --- | --- | --- |
| DIRECT CLOSURE | 30 | 35 | 0.401 |
| FULL-THICKNESS SKIN GRAFT | 14 | 7 | 0.139 |
| FLAP: LOCAL OR DISTANT | 6 | 8 | 0.774 |

E
| GRADE OF SURGEON | INCOMPLETE | COMPLETE | P-VALUE |
| --- | --- | --- | --- |
| SHO | 0 | 0 | NA |
| REGISTRAR | 22 | 22 | 1.000 |
| CLINICAL SPECIALIST | 10 | 13 | 0.635 |
| CONSULTANT | 18 | 14 | 0.520 |

| FOLLOW UP |  |
| --- | --- |
| OBSERVATOPN | 31 |
| RE-EXCISION | 11 |
| RADIOTHERAPY | 6 |
| LOST | 2 |

For incomplete excision tumours the methods of reconstruction following tumour resection were direct closure of the wound in 60% (30/50), 28% (14/50) using a full thickness skin graft and 12% (6/50) with a local or distant flap. In contrast, for complete excision tumours 70% (35/50) received direct closure of the wound, 14% (7/50) required a full thickness skin graft and 16% (8/50) underwent a local or distant flap. There was no statistically significant difference between the method of reconstruction between incomplete and complete excision tumours (Table D). Moreover, there was no significant difference between the seniority of surgeon (*p*=0.520) and the risk of incomplete excision and in both tumours a registrar performed the operation most frequently (Table E).

All patients with completely excised tumours were discharged following treatment and during the 12 month follow up period none (0/50) suffered disease recurrence. For incomplete excision tumours, 62% (31/50) were followed up by surveillance only, 22% (11/50) underwent a repeat excision, 12% (6/50) received post-operative radiotherapy and 4% (2/50) were lost to follow up (Table F).

## Discussion

Facial BCC represents a common and heterogeneous malignancy that demands detailed investigation to inform optimum management. The reported incomplete excision rate in our study was 10.96% within the range of 4 to 16.6% described in the literature^7,8^. Crucially, our focused investigation of facial BCC provides improved resolution to support the classification of high-risk tumours. We demonstrate that tumour histology is a significant factor influencing the likelihood of complete excision with mixed, infiltrative and morphoeic subtypes significantly increased in tumours with involved margins. Moreover, we show nodular variants are significantly reduced in incomplete excision tumours supporting this subset as more indolent with a less aggressive growth pattern. Although distinct subtypes are established in stratifying low and high risk tumours our results confirm that tumour histology in facial BCC impacts upon the success of surgical excision. In future, for more aggressive tumour variants this data suggests additional techniques such as Mohs micrographic surgery, intra-operative frozen section or a wider excision margin may be appropriate to maximize tumour clearance. The BAD guidelines for large or morphoeic tumours state that a 13-15mm peripheral margin will clear the tumor in >95% and a margin of at least 5mm is appropriate. This provides a guide for peripheral margins in high risk facial BCC.

Interestingly, our data also show that tumour location at the ala nasi and inner canthus is a significant factor influencing the likelihood of complete surgical excision. Previous studies have characterized high-risk anatomical areas as the ear, eyelid and temple^9^ and our results provide detail to highlight discrete regions of the face with the highest risk of involved margins. This observation supports high risk tumours positioned at areas of embryological fusion and theorized as lines of innate weakness in tissue architecture that allow tumour infiltration. One central challenge in treating facial BCC is maintaining the preservation of healthy tissue to optimize the aesthetic outcome whilst not jeopardizing adequate surgical margins. Indeed, this principle likely explains why tumours at the inner canthus and ala nasi are at higher risk of involved margins. Together, our results define tumour location and histology as significant factors stratifying high risk tumours in facial BCC.

In this study we show the deep margin (40%, 20/50) is more frequently involved as compared to the peripheral margin (28% 14/50), in line with previous results. This suggests the deep margin may be more demanding to judge macroscopically. Indeed, BAD guidelines align with this data and suggest that the deep margin is more difficult to define due to the dependence upon local anatomy and recommend excision extending to subcutaneous fat. In contrast, Farhi et al (2007) and Breuninger & Dietz (1991) showed no significant difference between involved margins and Bartos et al (2003) reported an incomplete excision rate of 10.3% with 8.6% positive lateral margins and 2.5% positive deep margins. It is beyond the scope of this study to define how each surgical margin correlates with the risk of recurrence and there appears to be conflicting results in the literature. Given the three-dimensional infiltration pattern of BCC, and the unpredictable nature of tissue invasion, it is challenging to explain how each surgical margin may relate to the overall likelihood of tumour clearance. Nevertheless, our result supports a differential involvement of deep and peripheral margin in facial BCC. Further work should aim to explore whether distinct clinical and pathological features correlate with the involvement of specific margins and how this could inform surgical planning.

Limitations to our study relate to relatively small sample size and this may have influenced the lack of statistically significant differences between variables such as grade of surgeon or method of reconstruction. One surprising finding was the lack of any disease recurrence in our cohort despite 62% (31/50) of tumours with involved margins being followed up with surveillance alone. This may however relate to the potential long latency period of recurrence and the relatively short follow up for tumours in our study (6-17 months). In future, long term follow up data of facial BCC will provide importance insights into outcomes. Finally, our study focused on conventional surgical excision in accordance with BAD guidelines however it would be interesting to see how different therapeutic methods, including Mohs micrographic surgery, relate to distinct clinical and pathological features of incompletely excised tumours.

## Conclusion

Basal cell carcinoma is the most common malignancy in humans and its potential for local invasion mandates a pragmatic approach to treatment. Although rare, incomplete excision of facial BCC requires careful consideration to optimize patient outcomes. Importantly, our data show tumour location and histology as key characteristics determining the success of tumour clearance and we provide a valuable new patient cohort of facial BCC. Our results highlight the deep margin as a crucial factor to be considered in high risk facial BCC and suggest a peripheral margin of at least 5mm should be used for these tumours. In future, we should aim to integrate clinical, pathological, and genomic data to build a robust facial cancer signature in BCC to enable more personalized treatment strategies.

## Data Availability

All data produced in the present work are contained in the manuscript

## References

1. Clark, C.M., Furniss, M. & Mackay-Wiggan, J.M. Basal cell carcinoma: an evidence-based treatment update. Am J Clin Dermatol 15, 197–216 (2014).

2. Montagna, E. & Lopes, O.S. Molecular basis of basal cell carcinoma. An Bras Dermatol 92, 517–520 (2017).

3. Griffiths, R.W., Suvarna, S.K. & Stone, J. Basal cell carcinoma histological clearance margins: an analysis of 1539 conventionally excised tumours. Wider still and deeper? Journal of Plastic, Reconstructive & Aesthetic Surgery 60, 41–47 (2007).

4. Levell, N.J., Igali, L., Wright, K.A., et al. Basal cell carcinoma epidemiology in the UK: the elephant in the room. Clin Exp Dermatol 38, 367–369 (2013).

5. Gulleth, Y., Goldberg, N., Silverman, R.P., et al. What is the best surgical margin for a Basal cell carcinoma: a meta-analysis of the literature. Plastic and reconstructive surgery 126, 1222–1231 (2010).

6. Telfer, N.R., Colver, G.B., Morton, C.A., et al. Guidelines for the management of basal cell carcinoma. Br J Dermatol 159, 35–48 (2008).

7. Mosterd, K., Krekels, G.A.M., Nieman, F.H., et al. Surgical excision versus Mohs’ micrographic surgery for primary and recurrent basal-cell carcinoma of the face: a prospective randomised controlled trial with 5-years’ follow-up. Lancet Oncol 9, 1149–1156 (2008).

8. Fernandes, J.D., de Lorenzo Messina, M.C., de Almeida Pimentel, E.R., et al. Presence of residual basal cell carcinoma in re-excised specimens is more probable when deep and lateral margins were positive. J Eur Acad Dermatol Venereol 22, 704–706 (2008).

9. Nagore, E., Grau, C., Molinero, J., et al. Positive margins in basal cell carcinoma: relationship to clinical features and recurrence risk. A retrospective study of 248 patients. J Eur Acad Dermatol Venereol 17, 167–170 (2003).

